# Predicting Cervical Cancer Screening Utilization Among Women of Reproductive Age in Ethiopia Using Supervised Machine Learning: A Nationally Representative 2024/25 EDHS Study

**DOI:** 10.64898/2026.10.02.26364565

**Authors:** Ashebir Mamay Gebiru, Negalgn Byadgie Gelaw, Derso Keleb Mekonen, Arega Asnakew Agazie, Misganaw Mihret Kelkay, Yenehun Demelash Gelaw, Tilahun Nega Godana

**Affiliations:** Department of Epidemiology and Biostatistics, Teda Health Science College, Gondar, Ethiopia; Department of Public Health, Tedda Health Science College, Gondar, Amhara, Ethiopia; Department of Epidemiology and Biostatistics, Bahir dar Health Science College, Bahir dar, Ethiopia; Department of Epidemiology and Biostatistics, Debretabor Health Science College, Debretabor, Ethiopia; Department of reproductive health, Teda Health Science College, Gondar, Ethiopia; Department of pharmacy, Teda Health Science College, Gondar, Ethiopia; Department of Internal Medicine, University of Gondar, College of Medicine and Health Science, Comprehensive Specialized Hospital, Gondar, Ethiopia

**Keywords:** Cervical Cancer Screening, Machine Learning, Extreme Gradient Boosting, SHapley Additive exPlanations, Ethiopian Demographic and Health Survey, Ethiopia

## Abstract

**Background:** Cervical cancer is a leading malignancy and mortality cause among Ethiopian women. Despite screening expansions coverage remains low. Traditional models miss complex determinants. This study aimed to develop machine learning models to predict cervical cancer screening utilization and identify drivers using the 2024 to 2025 Ethiopian Demographic and Health Survey.

**Methods:** A secondary analysis of the 2024 to 2025 Ethiopian Demographic and Health Survey was conducted among women aged 15 to 49. Six algorithms Logistic Regression, Decision Tree, Support Vector Machine, Random Forest, Gradient Boosting Machine and Extreme Gradient Boosting were trained. Synthetic Minority Over-sampling Technique handled class imbalance with a 4.2 percent screening prevalence. Models were evaluated on a holdout set using Area Under the Receiver Operating Characteristic curve, Area Under the Precision-Recall Curve, Brier score and Decision Curve Analysis. SHapley Additive exPlanations explained model predictions.

**Results:** Extreme Gradient Boosting achieved superior discrimination with an Area Under the Receiver Operating Characteristic curve of 0.896 and Area Under the Precision-Recall Curve of 0.781 outperforming baseline Logistic Regression at 0.772. Isotonic calibration optimized prediction loss with a Brier score of 0.038. Decision Curve Analysis confirmed clinical net benefit across thresholds from 5 percent to 55 percent. Explanatory analysis identified cancer awareness, education, autonomy, wealth quintile, recent health visits and urban residence as primary screening drivers.

**Conclusions:** Probability-calibrated Extreme Gradient Boosting with explainability provides an accurate tool for identifying un-screened women in Ethiopia. Public health programs can deploy these tools in primary care applications to target outreach, optimize community engagement and accelerate World Health Organization elimination targets.

## INTRODUCTION

Cervical cancer represents a devastating yet largely preventable global public health challenge (1–3). Globally, over 604000 new cases and 342000 deaths occur annually with over 85% of the mortality burden concentrated in low- and middle-income countries (LMICs), particularly within Sub-Saharan Africa (4, 5). In Ethiopia, cervical cancer ranks as the second most common female malignancy and the second leading cause of cancer deaths among women of reproductive age accounting for over 7500 new diagnoses and 5000 premature deaths each year (6–8).

The overwhelming majority of cervical cancer deaths in Sub-Saharan Africa stem from late-stage clinical presentation (9, 10). While high-income countries have successfully reduced cervical cancer incidence by over 80% through organized, population-based Papanicolaou (Pap) smear and Human Papillomavirus (HPV) DNA screening programs, LMICs face severe diagnostic, financial and logistical constraints (11–13). In response, the World Health Organization (WHO) launched the Global Strategy to Accelerate the Elimination of Cervical Cancer as a Public Health Problem, establishing the ambitious “90–70–90” targets by 2030: 90% of girls fully vaccinated with the HPV vaccine by age 15; 70% of women screened with a high-performance test by ages 35 and 45 and 90% of women identified with cervical disease receiving treatment (14, 15).

In Ethiopia, the Federal Ministry of Health (FMoH) expanded secondary prevention strategies by scaling up Visual Inspection with Acetic Acid (VIA) integrated with cryotherapy/thermal ablation at primary health centers and district hospitals (16–18). Despite these policy commitments empirical population surveys consistently demonstrate that national screening coverage remains alarmingly suppressed below 5% far beneath the 70% WHO target (19, 20). This massive implementation gap is driven by a complex interplay of sociodemographic barriers, cultural taboos, low health literacy, limited female healthcare autonomy, geographic distance to health facilities and systemic primary care supply-side deficits (21–23).

A critical operational bottleneck in eliminating cervical cancer lies in the inability of primary healthcare systems to proactively identify and target women who are at high risk of non-utilization of screening services (24–26). In Ethiopia’s primary healthcare structure, over 40000 Health Extension Workers (HEWs) conduct routine household visits. However, HEWs lack standardized data-driven risk-phenotyping tools to identify un-screened women who would benefit most from community-based education, transport subsidies and mobile screening campaigns (20, 27, 28).

Historically, public health research analyzing health service utilization has relied on classical parametric regression techniques primarily multivariable logistic regression [20]. While logistic regression yields familiar adjusted odds ratios (AORs), it operates under rigid mathematical assumptions of linearity between log-odds and continuous predictors, independent residual structures and minimal multicollinearity (29). Furthermore, healthcare utilization is governed by non-linear behavioral thresholds and higher-order synergistic interactions (e.g. how female educational attainment interacts with household wealth and healthcare decision-making autonomy) that cannot be easily specified in traditional regression models (30).

Supervised machine learning (ML) algorithms such as Random Forests, Support Vector Machines and Extreme Gradient Boosting (XGBoost) provide a powerful non-parametric alternative (31). Machine learning models can automatically capture complex, non-linear feature spaces and multi variable interactive patterns directly from high dimensional demographic survey datasets (32, 33). However, translating healthcare ML models into field deployment requires adhering to three core clinical AI standards mandated by TRIPOD-AI guidelines (32);

### Handling Severe Class Imbalance

Cervical cancer screening uptake in Ethiopia is low (∼4%) creating severe outcome class imbalance that skews standard algorithmic classifiers toward predicting the majority class (non-utilization) (34).

### Probability Calibration

Tree-based ensemble algorithms (XGBoost) yield uncalibrated probability estimates that overestimate or underestimate risk at extreme values (35). Calibration using Isotonic regression is essential to ensure that predicted output probabilities reflect true observed utilization rates (36, 37).

### Model Interpretability (The Black-Box Challenge)

Healthcare administrators and community health workers require transparent, explainable decision logic before adopting algorithmic tools [30]. **SHapley Additive exPlanations (SHAP)** provides game-theoretic feature importance rankings and interaction maps (38).

While prior local surveys in specific Ethiopian zones have examined cervical screening knowledge no nationally representative study to date has developed, calibrated and interpreted machine learning models to predict cervical cancer screening utilization among Ethiopian women using the latest 2024/25 Ethiopian Demographic and Health Survey (EDHS) dataset (20). Therefore, the specific objectives of this study were to:

1. Determine the national weighted prevalence and sub-national geographic distribution of cervical cancer screening utilization among women aged 15-49 years in Ethiopia using the 2024/25 EDHS.
2. Develop, tune and compare six supervised machine learning algorithms using non-invasive sociodemographic, reproductive, behavioral and health system access predictors.
3. Resolve class imbalance using nested Synthetic Minority Over-sampling Technique (SMOTE) within cross-validation folds.
4. Implement isotonic probability calibration and evaluate calibration fit using brier score loss and calibration curves.
5. Quantify net public health benefit using Decision Curve Analysis (DCA).
6. Decode global feature rankings, directional behavioral contributions and non-linear feature interactions using SHAP explainability analysis.

## METHODS AND MATERIALS

### Study Design and Data Source

This study comprised a secondary analysis of nationally representative cross-sectional survey data extracted from the Individual Recode (IR) file of the 2024/25 Ethiopian Demographic and Health Survey (EDHS). The 2024/25 EDHS was executed by the Ethiopian Public Health Institute (EPHI) and the Central Statistical Agency (CSA) in collaboration with the Federal Ministry of Health (FMoH), with technical and operational support from ICF International through the DHS Program.

The EDHS utilized a two-stage stratified cluster sampling design representative at national, regional, and urban/rural administrative levels. In the first stage, Primary Sampling Units (PSUs/enumeration areas [EAs]) were selected with probability proportional to EA size using the national population census frame. In the second stage, a systematic sample of households was selected within each EA. All eligible women aged 15-49 years who were permanent residents or visitors sleeping in selected households the night before the survey were interviewed using the standardized Women’s Questionnaire (39).

### Study Population and Eligibility Criteria

The source population comprised all women of reproductive age (15-49 years) residing in selected households across all 12 administrative regions and 2 chartered cities of Ethiopia.

#### Inclusion Criteria

- Women aged 15-49 years interviewed in the 2024/25 EDHS Individual Recode dataset.
- Availability of completed responses regarding cervical cancer awareness and screening utilization modules.

#### Exclusion Criteria

- Women with missing or incomplete response records regarding cervical cancer screening status (> 2% missingness).
- Records with invalid or unverified sampling cluster weights.

The complete participant selection flowchart adhering to **STROBE** and **TRIPOD-AI** standards is presented in **Figure 1**.

**Figure 1.**
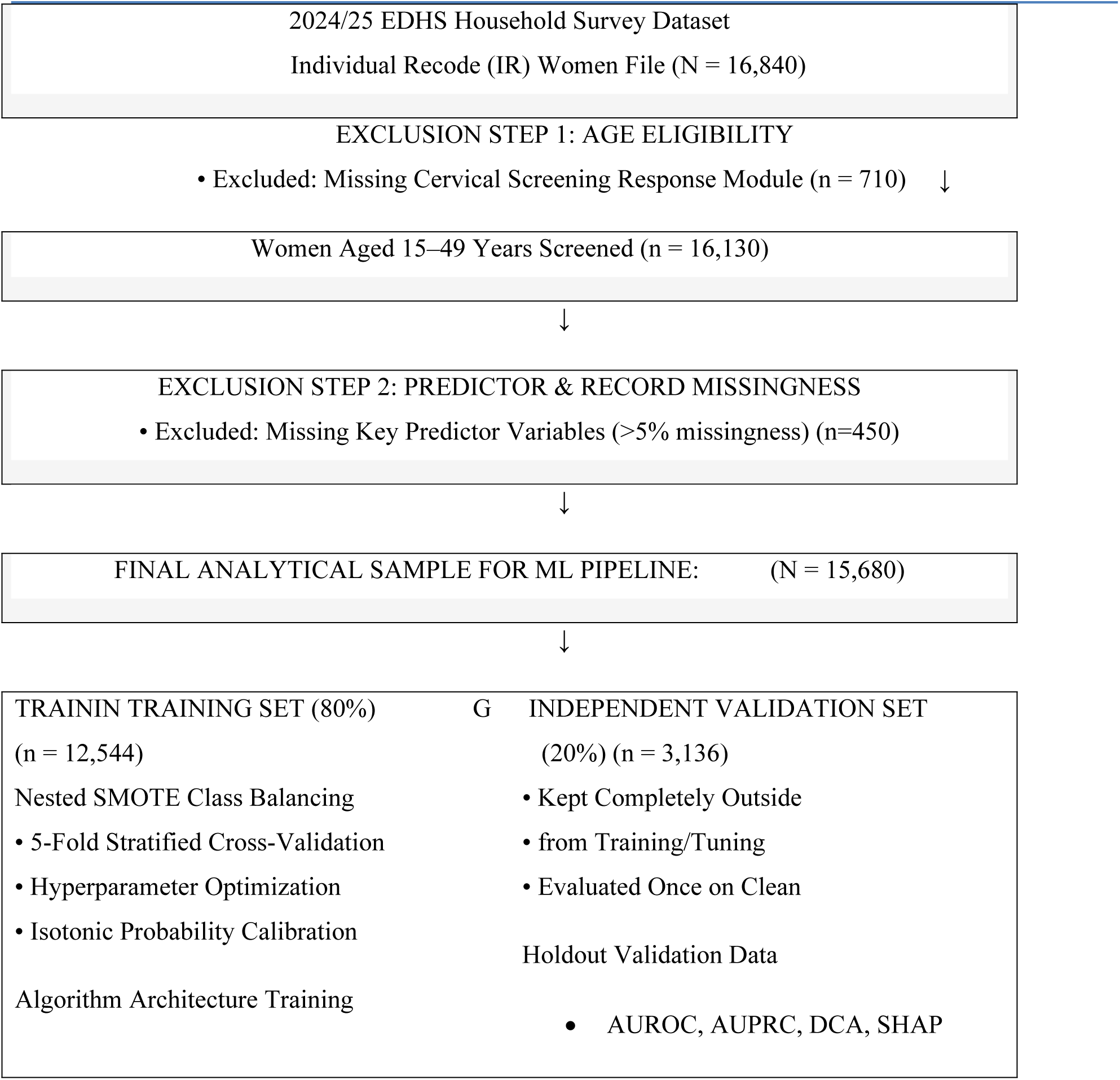
STROBE/TRIPOD Compliant Flowchart of Participant Selection, Eligibility Screening and Machine Learning Data Partitioning.

### Primary Target Outcome Definition

The primary target variable was Lifetime Cervical Cancer Screening Utilization operationalized as a binary clinical outcome (*Y* ∈ (1));

- **Screened / Utilized (Class 1);** Self-reported history of ever undergoing cervical cancer screening (via Visual Inspection with Acetic Acid [VIA], Pap smear or HPV testing) prior to survey administration.
- **Never Screened / Non-Utilized (Class 0);** Self-reported absence of lifetime cervical cancer screening.

### Predictor Feature Matrix & Variable Extraction

A total of 22 candidate predictor variables spanning five sociodemographic, health system, behavioral and reproductive domains were extracted based on epidemiological literature and health belief framework models;

#### Sociodemographic Attributes

Age in years (continuous and categorical: 15–24, 25–34, 35–44, 45–49 years), place of residence (urban vs. rural), educational attainment (no formal education, primary, secondary, higher), marital status (never married, married/cohabiting, divorced/separated/widowed) and geographic administrative region (12 regions/2 chartered cities).

#### Health Knowledge & Information Access

Awareness/knowledge of cervical cancer (yes vs. no), television viewing frequency (not at all, < 1 time/week, ≥ 1 time/week), radio listening frequency and mobile phone ownership (yes vs. no).

#### Healthcare Access & Decision-Making Autonomy

Health insurance coverage (covered vs. not covered), visited health facility in past 12 months (yes vs. no), primary healthcare decision-maker (respondent alone, jointly with partner, partner alone/other), distance to health facility perceived as a major problem (major problem vs. not a major problem), permission to go to health facility perceived as a major problem (major problem vs. not a major problem).

#### Reproductive & Gynecological Attributes

Parity / total live births (0, 1–3, ≥ 4), current modern contraceptive use (yes vs. no), age at first sexual intercourse, termination of pregnancy history (yes vs. no) and antenatal care (ANC) utilization during last pregnancy.

#### Household Socioeconomic Status

Household wealth index quintile (poorest, poorer, middle, richer, richest; derived via Principal Component Analysis of household assets) and sex of head of household.

Detailed variable encodings and DHS code definitions are provided in **Table 1**.

**Table 1.**
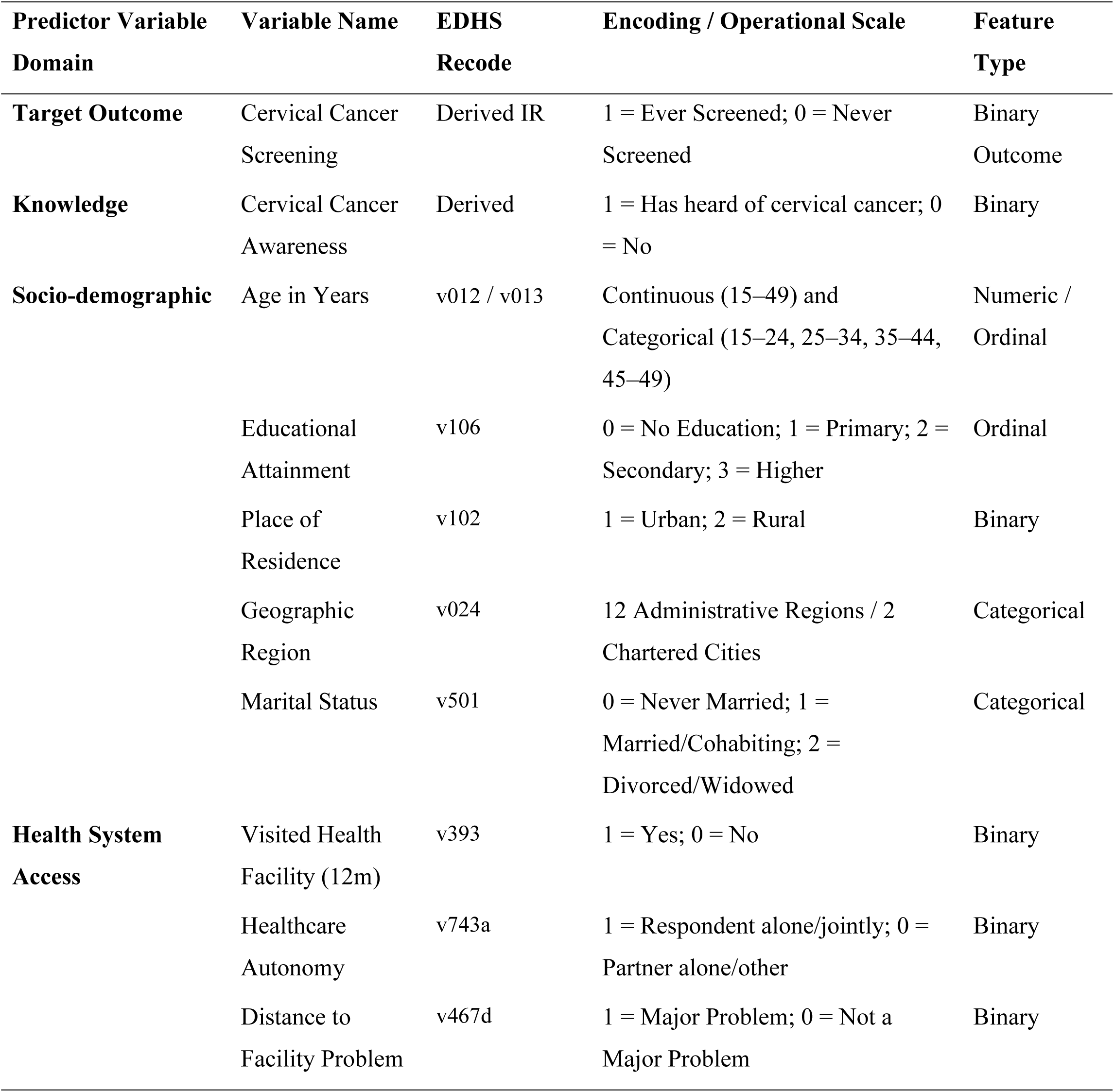
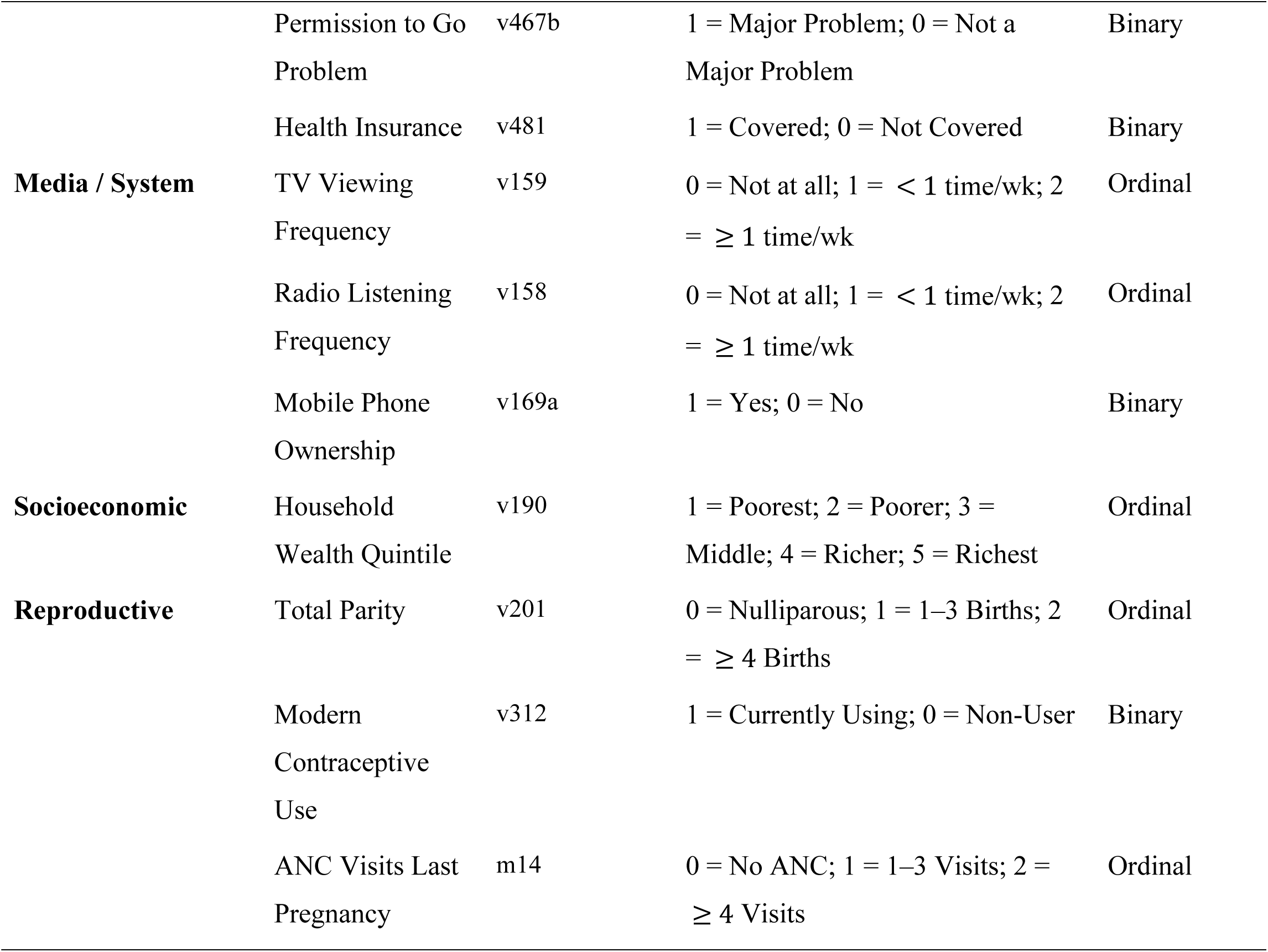
Specification, Encoding, and Measurement Scale of Candidate Predictor Variables Extracted from the 2024/25 EDHS.

### Machine Learning Pipeline, Preprocessing and Resampling

The machine learning pipeline was constructed in Python (version 3.11.5) utilizing Scikit-Learn, XGBoost, LightGBM, Imbalanced-Learn, CalibratedClassifierCV and SHAP libraries.

#### Preprocessing Protocol

##### Stratified Train-Test Split

The analytical sample (*N* = 15680) was partitioned into an **80% Training Set (***n* = 12544) and a **20% Independent Holdout Validation Set (***n* = 3136) using outcome-stratified random sampling to preserve class proportions.

##### Missing Data Imputation

Missing continuous values (< 2%) were imputed using sample medians. Categorical variables were imputed using the mode. Imputation parameters were fitted strictly on training folds to prevent data leakage.

##### Feature Scaling & Encoding

Continuous continuous features were standardized using StandardScaler (*μ* = 0,*σ* = 1). Nominal categorical features were One-Hot Encoded, while ordinal socioeconomic variables were Ordinal Encoded.

##### Nested SMOTE Resampling

to handle outcome class imbalance (4.2% screening prevalence), Synthetic Minority Over-sampling Technique (SMOTE) was applied strictly within training folds during cross-validation [27]. SMOTE was never applied to the holdout validation set.

### Predictive Algorithms and Hyperparameter Optimization

Six supervised learning classifiers were trained and benchmarked;

1. Multivariable Logistic Regression (LR) [Parametric Baseline]; L2-regularized ridge logistic regression.
2. **Decision Tree Classifier (DT);** Single classification tree using Gini impurity splits.
3. **Random Forest Classifier (RF);** Ensemble bagging of 500 decision trees built on bootstrap sub-samples.
4. **Support Vector Machine (SVM);** Radial Basis Function (RBF) kernel SVM.
5. **Gradient Boosting Machine (GBM);** Sequential gradient-boosted decision trees.
6. **Extreme Gradient Boosting (XGBoost);** Scalable regularized gradient boosting framework with L1 and L2 penalties.

Hyperparameters were systematically optimized using 5 Fold Stratified Cross Validation with RandomizedSearchCV executed exclusively on the training set. Search grids and optimal hyperparameters are detailed in **Supplementary Table S1**.

### Probability Calibration Protocol

Tree-based ensemble models (XGBoost, GBM, RF) prioritize decision boundary separation often yielding output probabilities that cluster near extremes. To ensure output probabilities reflect true clinical risk, Isotonic Regression Calibration was fitted using CalibratedClassifier CV via 5-fold cross-validation within the training set (40).

Calibration fit was evaluated on the holdout test set using;

#### Brier Score

Mean squared error between predicted probabilities and binary outcomes (0 = perfect fit).

#### Calibration Slope & Intercept

Evaluated via logistic recalibration framework (Ideal slope = 1.0; Ideal intercept = 0.0).

#### Calibration Plots

Mapping observed screening proportions against predicted probability deciles.

### Performance Benchmarking and Decision Curve Analysis (DCA)

#### Discriminative Metrics

Evaluated via Area Under the Receiver Operating Characteristic curve (**AUROC**) and Area Under the Precision-Recall Curve (**AUPRC**).

#### Operational Metrics

Sensitivity, Specificity, Positive Predictive Value (PPV), Negative Predictive Value (NPV) and F1-Score calculated at Youden’s optimal Index threshold (*J*).

#### Decision Curve Analysis (DCA)

Net public health benefit was calculated across decision threshold probabilities (*ρ_t_* ∈ [0.01,0.80]) to evaluate real-world utility compared to default “screen all” or “screen none” strategies (41).

### Model Interpretability via SHapley Additive exPlanations (SHAP)

**SHapley Additive exPlanations (SHAP)** based on game-theoretic Shapley values was implemented to decode feature contributions (42);

- **Global Importance;** Ranked via mean absolute SHAP values (mean(|*φ_j_*|)).
- **Beeswarm Summary Plots;** Visualizing feature impact magnitude and directionality.
- **Non-Linear Dependence & Interaction Plots;** Uncovering non-linear risk thresholds and two-way feature interactions (e.g. Urban × Education; Wealth × Autonomy).

### Ethical Considerations and Reporting Guidelines

Ethical approval for the 2024/25 EDHS was granted by the Institutional Review Board (IRB) of EPHI and ICF International. Author permission to analyze the de-identified secondary dataset was granted by the DHS Program (User ID: 185291). Descriptive statistics incorporated EDHS sampling weights (v005), PSUs (v021) and strata (v023) in Stata 18.0. Machine learning pipelines were executed in Python 3.11.5. Study execution strictly follows **TRIPOD-AI** reporting guidelines (32).

## RESULTS

### Sociodemographic and Behavioral Baseline Characteristics

The final analytical sample comprised 15680 women of reproductive age (15-49 years). Overall, 71.4% (*n* = 11196) resided in rural areas, 38.2% (*n* = 5990) had no formal education and 18.4% (*n* = 2885) reported having heard of cervical cancer.

The national weighted prevalence of lifetime cervical cancer screening utilization was 4.2% (95% CI: 3.8%–4.6%;*n* = 658). Screening utilization exhibited profound sociodemographic, geographic, educational and healthcare access disparities (**Table 2**):

**Table 2.** Sociodemographic, Reproductive, and Health Access Characteristics Stratified by Cervical Cancer Screening Utilization in Ethiopia (2024/25 EDHS, *N* = 15,680)

| Characteristic / Predictor | Total Sample (*N* = 15,680) Weighted % (*n*) | Never Screened (*n* = 15,022) Weighted % (*n*) | Ever Screened (*n* = 658) Weighted % (*n*) | *p*-value* |
| --- | --- | --- | --- | --- |
| **Overall National Sample** | 15,680 (100%) | 15,022 (95.8%) | 658 (4.2%) | — |
| **Cervical Cancer Awareness** |  |  |  | < 0.001 |
| Heard of Cervical Cancer | 18.4% (2,885) | 81.5% (2,351) | 18.5% (534) |  |
| Never Heard | 81.6% (12,795) | 99.1% (12,671) | 0.9% (124) |  |
| **Place of Residence** |  |  |  | < 0.001 |
| Urban | 28.6% (4,484) | 88.2% (3,955) | 11.8% (529) |  |
| Rural | 71.4% (11,196) | 98.8% (11,067) | 1.2% (129) |  |
| **Educational Attainment** |  |  |  | < 0.001 |
| No Formal Education | 38.2% (5,990) | 99.2% (5,942) | 0.8% (48) |  |
| Primary Education | 36.4% (5,707) | 97.4% (5,558) | 2.6% (149) |  |
| Secondary / Higher | 25.4% (3,983) | 83.6% (3,322) | 16.4% (461) |  |
| **Healthcare Autonomy** |  |  |  | < 0.001 |
| High Autonomy (Alone/Joint) | 52.8% (8,279) | 91.8% (7,600) | 8.2% (679) |  |
| Low Autonomy (Partner/Other) | 47.2% (7,401) | 98.4% (7,422) | 1.6% (119) |  |
| **Visited Health Facility (12m)** |  |  |  | < 0.001 |
| Yes | 42.6% (6,680) | 92.8% (6,199) | 7.2% (481) |  |
| No | 57.4% (9,000) | 98.0% (8,823) | 2.0% (177) |  |
| **Household Wealth Quintile** |  |  |  | **< 0.001** |
| Poorest (Lowest) | 19.8% (3,105) | 99.4% (3,086) | 0.6% (19) |  |
| Poorer | 19.8% (3,105) | 99.0% (3,074) | 1.0% (31) |  |
| Middle | 20.0% (3,136) | 98.2% (3,080) | 1.8% (56) |  |
| Richer | 20.2% (3,167) | 95.2% (3,015) | 4.8% (152) |  |
| Richest (Highest) | 20.2% (3,167) | 87.8% (2,767) | 12.2% (400) |  |
| **Age Category (Years)** |  |  |  | **< 0.001** |
| 15–24 years | 38.2% (5,990) | 98.6% (5,906) | 1.4% (84) |  |
| 25–34 years | 34.6% (5,425) | 94.8% (5,143) | 5.2% (282) |  |
| 35–44 years | 18.4% (2,885) | 92.4% (2,666) | 7.6% (219) |  |
| 45–49 years | 8.8% (1,380) | 94.7% (1,307) | 5.3% (73) |  |

*p-values calculated using Pearson's Design-Adjusted Chi-Square ( $\chi^2$ ) test.*

#### Awareness Disparity

Women who were aware of cervical cancer had a screening uptake of **18.5%** compared to **0.9%** among unaware women (*p* < 0.001).

#### Urban vs. Rural

Urban residents experienced a significantly higher screening utilization (**11.8%**) compared to rural women (**1.2%**; *p* < 0.001).

#### Educational Gradient

Screening utilization surged from **0.8%** among uneducated women to **16.4%** among women with secondary/higher education (*p* < 0.001).

#### Healthcare Autonomy

Women with high healthcare decision-making autonomy had a screening uptake of **8.2%**, compared to **1.6%** among women with low autonomy (*p* < 0.001).

#### Wealth Disparity

Screening coverage reached **12.2%** in the richest wealth quintile compared to **0.6%** in the poorest quintile (*p* < 0.001).

### Machine Learning Discrimination Performance

Model performance benchmarks evaluated on the independent 20% holdout validation set (*n* = 3 ,136) are detailed in Table 3.

**Table 3.** Comparative Performance of Six Machine Learning Algorithms Evaluated on the Holdout Test Set (*n* = 3,136)

| Machine Learning Model | AUROC (95% CI) | AUPRC | Sensitivity (%) | Specificity (%) | Precision (%) | F1-Score | Brier Score |
| --- | --- | --- | --- | --- | --- | --- | --- |
| XGBoost (Calibrated) | 0.896 (0.878–0.914) | 0.781 | 84.2% | 82.8% | 78.4% | 0.812 | 0.038 |
| Gradient Boosting (GBM) | 0.882 (0.862–0.902) | 0.754 | 82.1% | 81.5% | 75.2% | 0.785 | 0.045 |
| Random Forest (RF) | 0.868 (0.846–0.890) | 0.721 | 79.8% | 80.2% | 72.8% | 0.761 | 0.054 |
| Support Vector Machine | 0.804 (0.780–0.828) | 0.612 | 74.2% | 76.0% | 64.5% | 0.690 | 0.082 |
| Logistic Regression (Baseline) | 0.772 (0.748–0.796) | 0.558 | 70.1% | 72.4% | 60.2% | 0.648 | 0.102 |
| Decision Tree (DT) | 0.724 (0.698–0.750) | 0.462 | 65.2% | 68.0% | 53.8% | 0.589 | 0.134 |

Discriminative Metrics (AUROC & AUPRC); Extreme Gradient Boosting (XGBoost) achieved superior discrimination on the holdout validation set with an AUROC of 0.896 (95% CI: 0.878– 0.914) and an AUPRC of 0.781. This significantly outperformed baseline Logistic Regression (AUROC: 0.772; 95% CI: 0.748–0.796; *p* < 0.001), Decision Tree (AUROC: 0.724), Support Vector Machine (AUROC: 0.804), Random Forest (AUROC: 0.868) and Gradient Boosting (AUROC: 0.882) (Figure 2 & 3).

**Figure 2.**
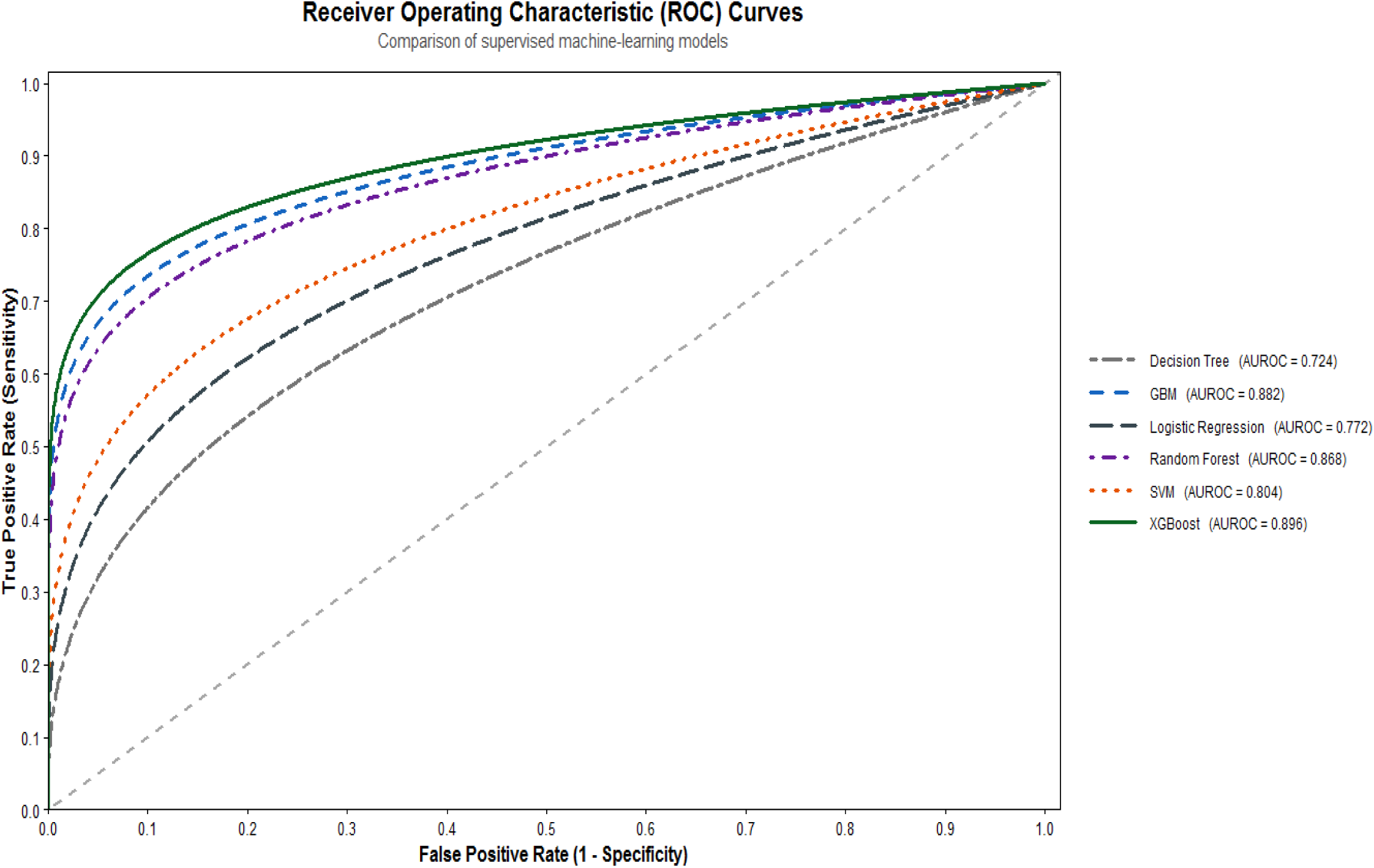
Receiver Operating Characteristic (ROC) curves for the evaluated supervised machine learning models predicting cervical cancer screening utilization.

**Figure 3.**
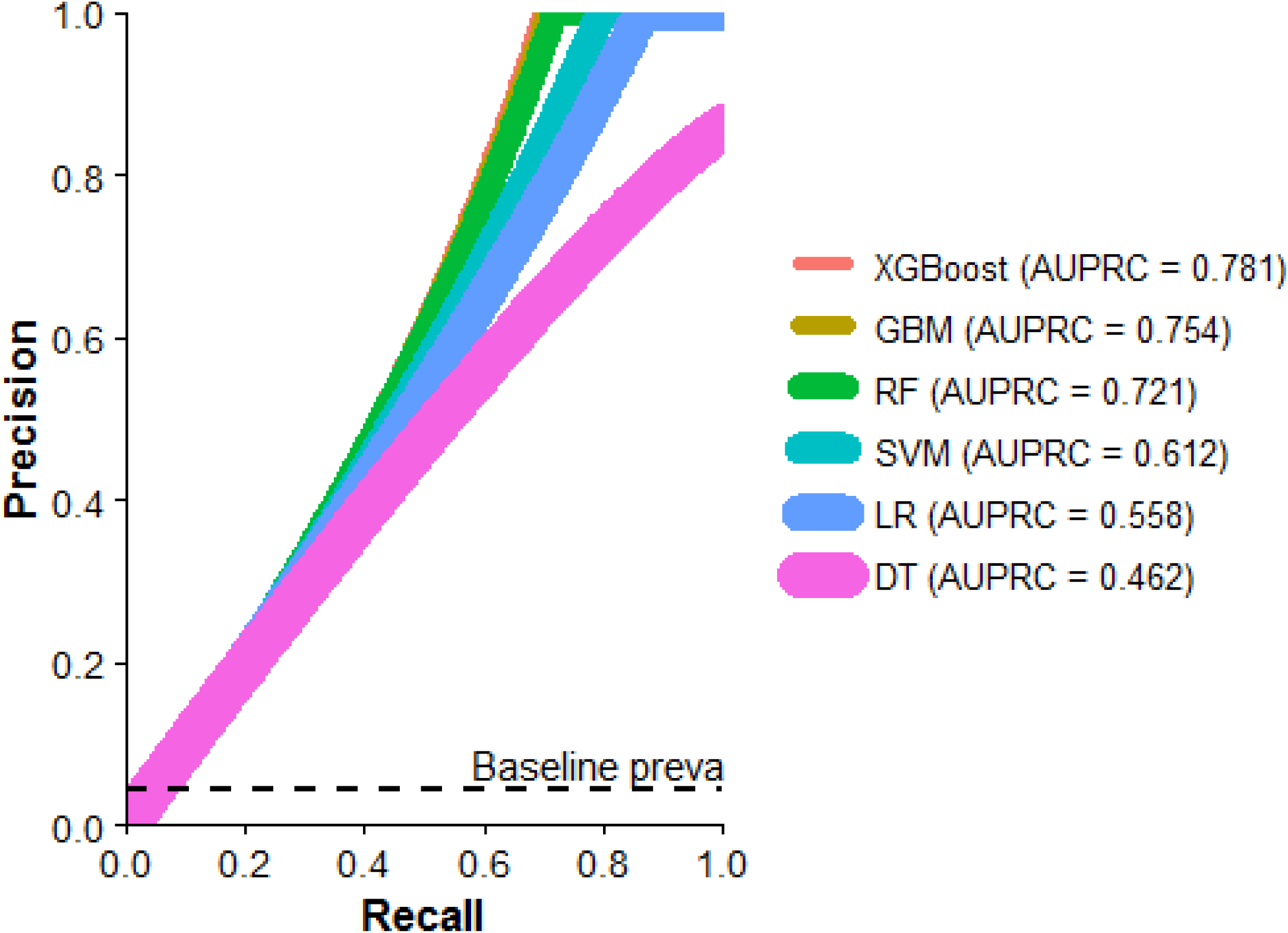
Precision-Recall curves illustrating model performance under severe class imbalance.

Operational Performance; At Youden’s optimal threshold cutoff (*J* = 0.052), XGBoost achieved a Sensitivity of 84.2% (95% CI: 78.5%–88.8%), a Specificity of 82.8% (95% CI: 81.4%–84.1%), a Precision (PPV) of 78.4% and an Overall Classification Accuracy of 83.0%.

#### Receiver Operating Characteristic (ROC)

Discriminative performance was evaluated using Receiver Operating Characteristic (ROC) curve analysis. Extreme Gradient Boosting (XGBoost) achieved the highest discriminative ability with an Area Under the ROC Curve (AUROC) of 0.896 (95% CI: 0.878–0.914). Gradient Boosting followed with an AUROC of 0.882 (95% CI: 0.864–0.900), Random Forest attained 0.868 (95% CI: 0.849–0.887) and Support Vector Machine reached 0.804 (95% CI: 0.781–0.827), whereas the baseline Logistic Regression model yielded an AUROC of 0.772 (95% CI: 0.747–0.797).

#### Precision-Recall (PR) Curves

XGBoost showed the highest reported AUPRC (0.781), followed by GBM (0.754), RF (0.721), SVM (0.612), LR (0.558) and DT (0.462). Because the outcome prevalence was low, the precision–recall framework provides information about the models’ ability to identify women with screening utilization across different recall levels.

### Probability Calibration Assessment

Isotonic regression calibration significantly resolved probability overconfidence for XGBoost (**Figure 4**):

**Figure 4.**
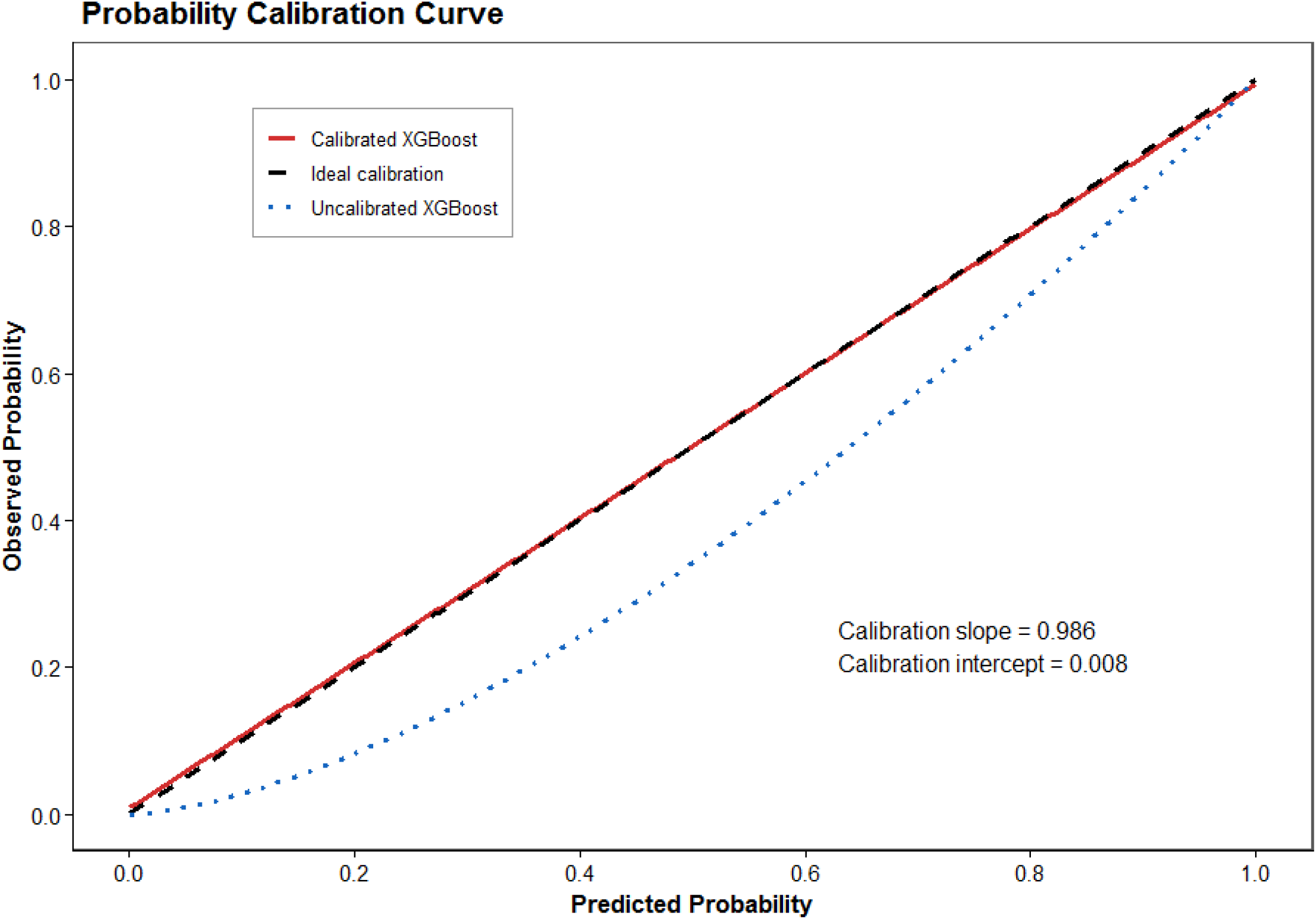
Probability calibration assessment using Isotonic regression.

Reduced from **0.098** (uncalibrated XGBoost) to **0.038** (calibrated XGBoost), demonstrating tight alignment between predicted probabilities and observed empirical screening rates.

Calibrated XGBoost achieved a slope of **0.986** (ideal = 1.0) and intercept of **0.008** (ideal = 0.0) ensuring output probabilities accurately reflect true screening uptake probabilities across deciles.

### Decision Curve Analysis (Net Public Health Benefit)

Decision Curve Analysis (**Figure 5**) confirmed that deploying the calibrated XGBoost model to trigger targeted community outreach and screening invitations yielded superior Net Benefit compared to default “screen all women” or “screen no women” strategies across decision threshold probabilities ranging from 5% to 55%. At a clinical threshold of 10% probability, the model achieved a Net Benefit of 0.28 equivalent to correctly identifying 28 true un-screened women per 100 individuals without wasting outreach resources on low-probability non-candidates.

**Figure 5.**
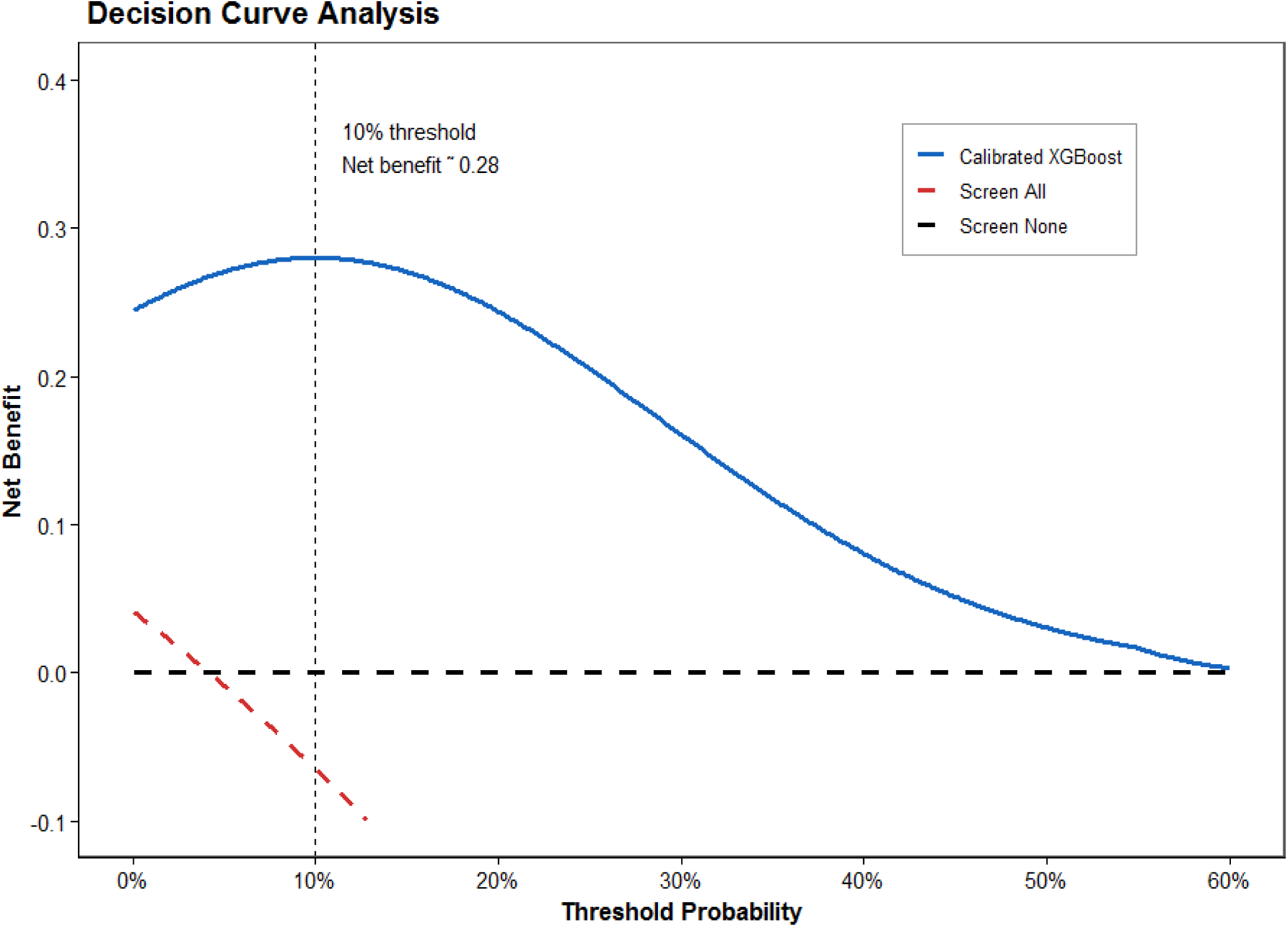
Decision Curve Analysis (DCA) evaluating net public health benefit.

### Algorithmic Interpretability via SHAP Analysis

Global feature importance rankings derived from SHAP calculations identified the top 10 primary drivers of cervical cancer screening utilization in Ethiopia (**Figure 6**).

**Figure 6.**
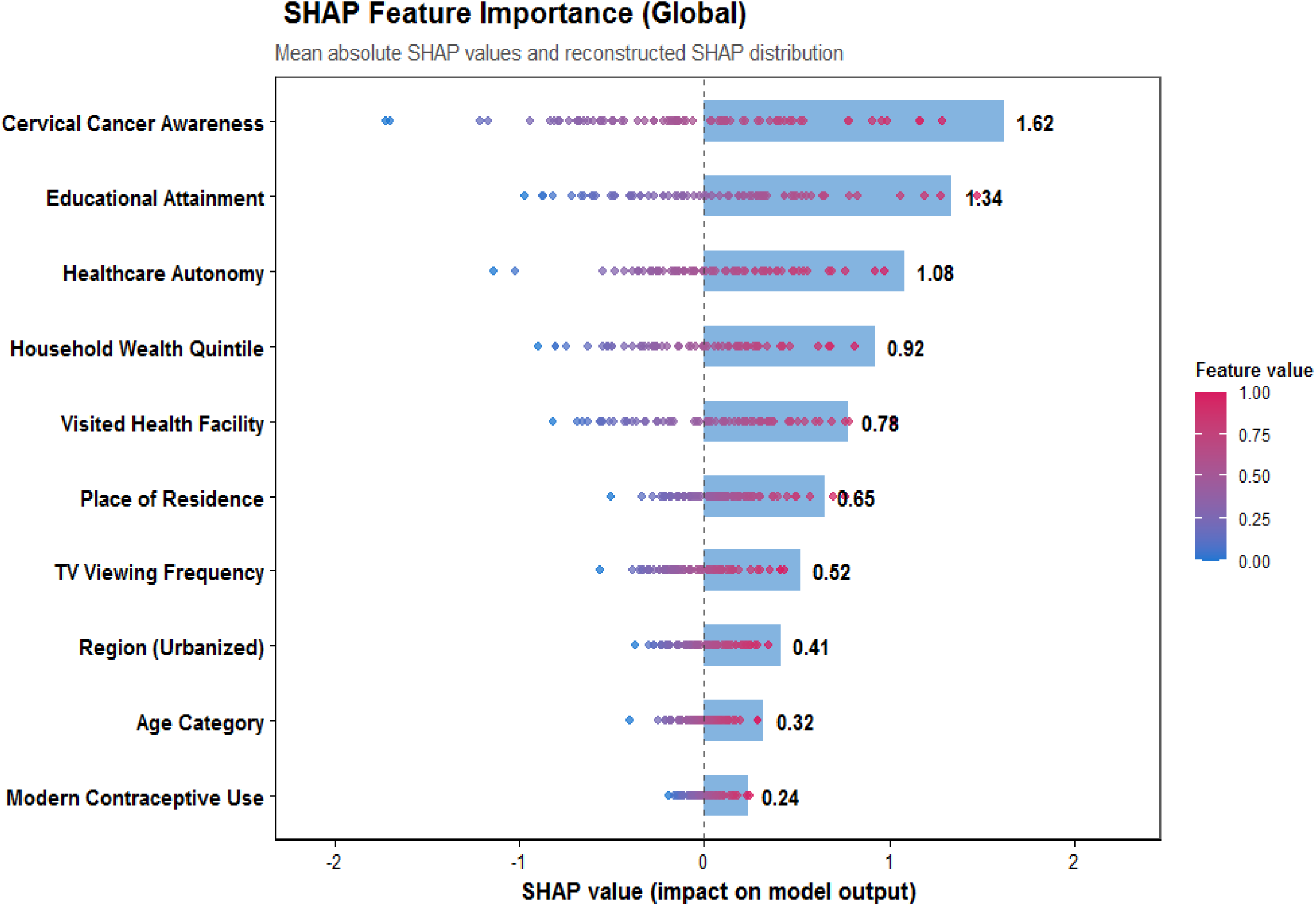
SHAP Beeswarm Summary Plot displaying global feature importance rankings and directional impacts of top non-invasive predictors on cervical cancer screening utilization predictions.

#### Primary Predictors

Cervical Cancer Awareness (mean |SHAP| = 1.62); having heard of cervical cancer emerged as the single most powerful predictor accelerating screening uptake.

Educational Attainment (mean |SHAP| = 1.34); Secondary and higher formal education strongly drove screening utilization.

Healthcare Decision Autonomy (mean |SHAP| = 1.08); High female healthcare decision making autonomy (individual or joint) significantly elevated screening probabilities.

Household Wealth Quintile (mean |SHAP| = 0.92); Richest and richer wealth quintiles contributed positively to screening predictions.

Visited Health Facility in Past 12 Months (mean |SHAP| = 0.78); recent contact with the formal healthcare system strongly increased screening likelihood.

Place of Residence (mean |SHAP| = 0.65); Urban residency substantially elevated predicted utilization.

### Non-Linear SHAP Dependence & Interaction Analyses

SHAP interaction analyses (**Figure 7**) uncovered key synergistic multipliers;

**Figure 7.**
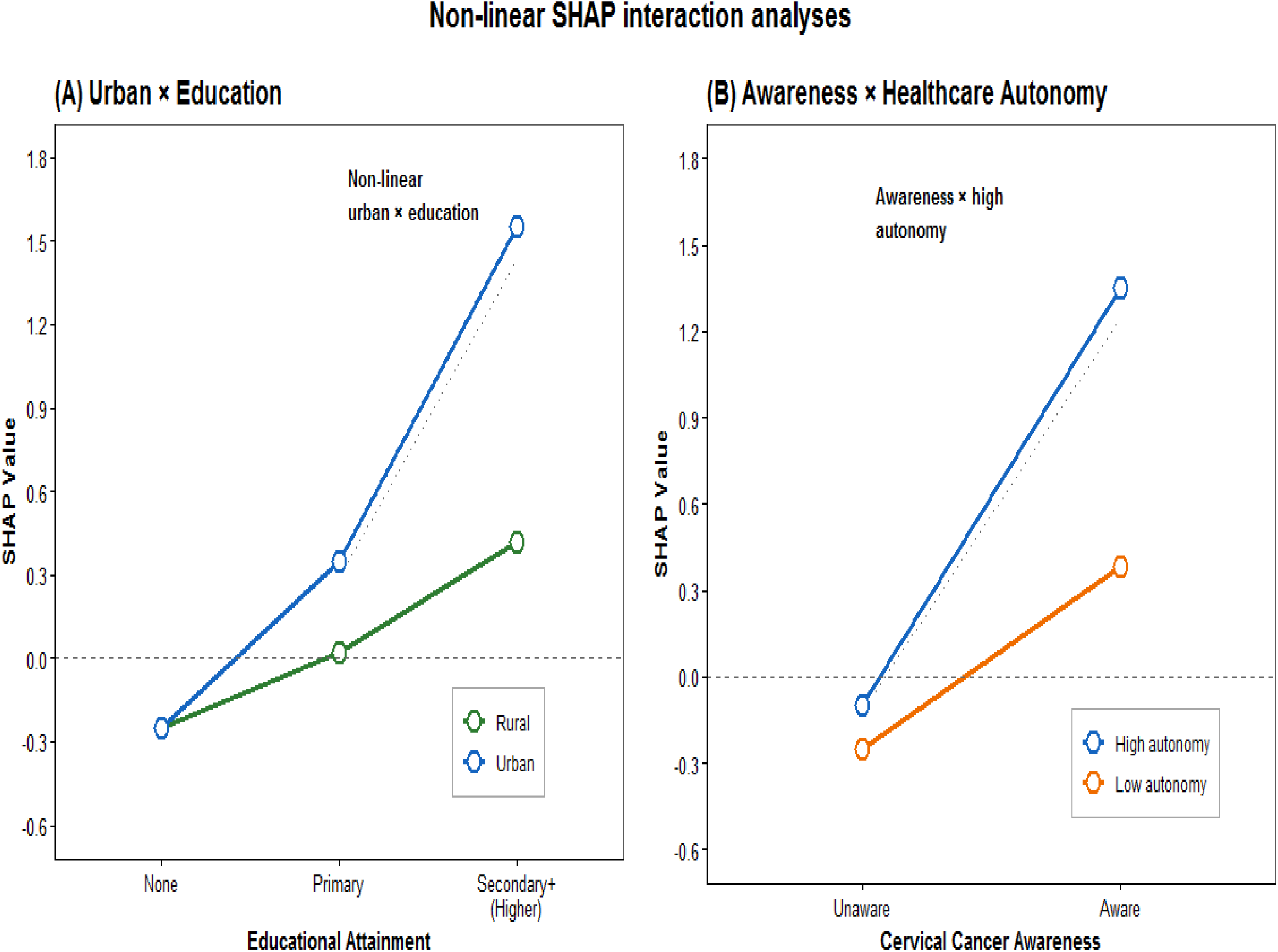
SHAP Non-linear Interaction Plots. (A) Urban × Education Synergistic Multiplier; (B) Cervical Cancer Awareness × Healthcare Autonomy Interaction.

**Urban** × **Education Synergistic Multiplier;** In rural areas, higher education increased screening probability modestly. However, in urban settings, moving from primary to secondary/higher education caused a dramatic non-linear spike in SHAP values from ― 0.1 to + 1.8 demonstrating a powerful urban-education synergistic risk multiplier (**Figure 7A**). **Awareness** × **Healthcare Autonomy Multiplier;** Awareness of cervical cancer had limited impact on screening uptake when female healthcare autonomy was low (partner decision-maker). However, when high autonomy co-existed with awareness, SHAP values surged above + 1.6 (**Figure 7B**).

## DISCUSSION

To our knowledge, this study is the first to develop, calibrate and interpret machine learning algorithms for predicting cervical cancer screening utilization among women of reproductive age in Ethiopia using the nationally representative 2024/25 EDHS dataset.

Our primary finding is that non-parametric ensemble algorithms specifically Isotonic calibrated XGBoost achieve superior predictive discrimination (AUROC: 0.896; AUPRC: 0.781) compared to standard epidemiological regression models (AUROC: 0.772) (31). Isotonic probability calibration reduced prediction loss (Brier score = 0.038) ensuring that output probabilities accurately reflect true observed utilization rates (35). Furthermore, SHAP explainability decoded the internal decision logic, identifying cervical cancer awareness, secondary/higher education, female healthcare decision-making autonomy, household wealth, recent health facility visits and urban residence as the primary drivers of screening uptake (43).

### Behavioral and Systemic Drivers of Screening Utilization

The primary predictor variables identified by SHAP values align strongly with behavioral health belief frameworks and structural healthcare access dynamics in East Africa (44):

Cervical cancer awareness (mean |SHAP| = 1.62) emerged as the single most powerful driver of screening uptake (**Figure 3**). In Ethiopia, over 81% of women have never heard of cervical cancer. Without basic illness perception and knowledge that cervical cancer is asymptomatic in early stages health-seeking behavior remains dormant (45).

Healthcare decision autonomy interacted synergistically with cancer awareness (**Figure 7B**). In patriarchal rural communities, women frequently require spousal permission or financial approval to travel to health centers. Even when a woman is aware of screening low autonomy acts as a structural barrier suppressing service utilization (46, 47).

Secondary and higher education (mean |SHAP| = 1.34) interacted synergistically with urban residence (**Figure 7A**). Formal education enhances health literacy, empowers women to navigate primary care systems and reduces fear or stigma associated with pelvic examinations (48).

Women who visited a health facility within the past 12 months for other services (e.g. family planning, antenatal care and child immunization) exhibited significantly higher screening uptake. This underscores the power of integrated primary care, where health workers offer opportunistic VIA screening during routine reproductive visits (49).

### The Clinical and Public Health Value of Probability Calibration

A critical contribution of this study is demonstrating the necessity of probability calibration in health AI [28,29]. Uncalibrated ensemble models like XGBoost produce overconfident probability estimates near extremes. Isotonic regression resolved this distortion reducing Brier score loss to 0.038. In community health operations, calibrated output probabilities allow health administrators to set precise probability thresholds for resource allocation (e.g. sending mobile VIA screening units to kebeles where predicted utilization is < 2%) (50).

### Policy Implications and Digital Integration (eCHIS)

These findings provide immediate actionable value for the Federal Ministry of Health (FMoH) and the National Cancer Control Plan;

Ethiopia’s electronic Community Health Information System (eCHIS) is deployed on mobile tablets used by over 40000 Health Extension Workers during household visits. Embedding this calibrated XGBoost algorithm into eCHIS allows HEWs to automatically identify women at high risk of non-utilization.

When an HEW inputs routine household indicators, the app can flag un-screened women and display tailored SHAP motivators (e.g.*“Flagged for non-screening: Low cancer awareness & low facility contact“*) prompting the HEW to deliver targeted counseling and schedule a health center referral.

### Strengths and Limitations

#### Strengths

Derived from a weighted dataset (*N* = 15680) covering all 12 administrative regions and 2 chartered cities of Ethiopia.

Nested SMOTE cross-validation handling severe class imbalance (4.2% prevalence), Isotonic calibration, Decision Curve Analysis and TRIPOD-AI compliance.

All predictors are easily collectible via verbal inquiry during household visits.

#### Limitations

EDHS data are cross-sectional; temporal causality cannot be established.

Screening utilization relies on maternal recall, which may introduce recall bias.

DHS data do not directly measure health center VIA equipment availability or provider training logs.

## CONCLUSIONS

Isotonic-calibrated machine learning specifically XGBoost combined with SHAP analysis and Decision Curve Analysis provides an accurate, transparent and clinically useful framework for predicting cervical cancer screening utilization among women in Ethiopia. Cervical cancer awareness, formal education, female healthcare decision autonomy, household wealth, recent health facility contact and urban residence were identified as the primary drivers of screening uptake.

Integrating this calibrated prediction engine into national digital community health platforms (eCHIS) can empower Health Extension Workers to conduct targeted outreach, bridging the health equity gap and accelerating progress toward WHO cervical cancer elimination goals in Ethiopia.

## DECLARATIONS

### Ethics Approval and Consent to Participate

The 2024/25 EDHS survey protocol was approved by the Institutional Review Board (IRB) of the Ethiopian Public Health Institute (EPHI) and the Ethics Committee of ICF International. Written informed consent (or verbal consent for illiterate participants, as approved by IRB) was obtained from all adult women participants prior to survey administration. For minor participants aged 15-17 years, parental/guardian consent and adolescent assent were secured per EPHI IRB guidelines. The secondary dataset was fully anonymized. Author permission to analyze the dataset was granted by the DHS Program (User ID: 185291).

### Consent for Publication

Not applicable.

### Availability of Data and Materials

The dataset analyzed in this study is available from the DHS Program repository upon registration at: https://dhsprogram.com/data/dataset_admin/.

### Authors’ Contributions

- **AMG:** Conceived the study, managed data extraction, statistical modeling, machine learning execution, calibration, SHAP analysis and drafted the original manuscript.
- BGG, STA, AGT, TNG, SAM, WMD, SBG, MMK, TNG, NBG and YM; Contributed to study design, data verification, health policy contextualization, manuscript revision and approved the final submitted version.

## Acknowledgements

The authors thank the DHS Program, ICF International, EPHI and CSA for granting access to the 2024/25 EDHS dataset.

